# Responsible AI in Action: Planning through Implementation of a Mortality Model for Palliative Care

**DOI:** 10.1101/2025.08.06.25333148

**Authors:** Claudia Nau, Susan E. Wang, Mina Habib, Haoyuan Zhong, Lori Viveros, Sergio Mendoza Sida, Kathleen Mulvaney, Kevin Durr, Bing Han, Beth Creekmur, Scott D. Halpern, Katherine R. Courtright, Janet S. Lee, Huong Q. Nguyen

**Affiliations:** Kaiser Permanente Southern California, Department of Research and Evaluation, Pasadena, CA; Kaiser Permanente J. Tyson School of Medicine, Pasadena, CA; The Permanente Federation, Oakland, CA; Southern California Permanente Group. Los Angeles, CA; Department of Geriatrics and Palliative Care, Kaiser Permanente Southern California, Pasadena, CA; Southern California Permanente Medical Group, Woodland Hills, CA; Epic Systems Corporation, Verona, WI; Systems Solutions & Deployment, Southern California Permanente Medical Group, Pasadena, CA; Palliative and Advanced Illness Research (PAIR) Center, University of Pennsylvania, Philadelphia, PA; Department of Medicine, University of Pennsylvania, Philadelphia, PA; Department of Medical Ethics and Health Policy, University of Pennsylvania, Philadelphia, PA; Center for Health Incentives and Behavioral Economics, University of Pennsylvania, Philadelphia, PA

## Abstract

**Importance:** Interest in the use of prediction models to support referrals to palliative care is surging. Few high-performing models have been developed, implemented, and pilot-tested following responsible artificial intelligence (AI) principles and transparent reporting guidelines.

**Objective:** Collaborate with physicians and clinical informaticists to plan, develop, validate, implement, and pilot-test a high-performing, statistically fair prediction model—the Serious Illness Clinical Indicator (SICLI)—to support palliative care referrals following responsible AI principles.

**Design:** Cohort study of hospitalizations occurring from January 1st, 2022, to December 31st, 2022.

**Setting:** Kaiser Permanente Southern California

**Participants:** Patients 18+ years, not admitted to maternity or psychiatry departments and hospitalized for at least 36 hours.

**Exposures:** We augmented a commercial prediction model, the Epic End-of-Life-Care Index (EOLCI), by adding locally available predictors, including co-morbidity and laboratory acuity risk scores, nursing flow sheet and health care utilization data.

**Main Outcomes and Measures:** We predicted 12-month mortality risk using a split-sample design, least absolute shrinkage, and selection operator (LASSO) using logistic regression, and five-fold cross-validation. Performance was assessed via the Area Under the Receiver Operating Curve (AUC) and calibration plots. The final model was chosen by clinicians and informaticists for its balance of performance and parsimony. Responsible AI principles guided each development step.

**Results:** Twelve-month mortality rate among 133,043 hospitalizations of at least 36 hours was 23%. Patients who died were older (76±13 vs. 64±17 years) and had higher co-morbidity burden (Charlson, 4.49±3.14 vs. 2.06±2.44). SICLI was very well calibrated and outperformed the EOLCI (AUC of 0.87 (95% CI:0.86, 0.87) vs. 0.81, (95% CI: (0.80, 0.81)). SICLI’s high and very-high risk group (probability cut-off 0.6-<95 and >=0.95), produced positive predictive values (PPVs) of 72.0% and 94.0%, respectively. SICLI was implemented in KPSC’s Electronic Health Record. Post-implementation validation by a palliativist against the 12-month surprise question via review of 25 charts yielded a PPV of 100% and 80% for the very-high and high-risk groups. We report on model documentation, socialization, and governance.

**Conclusion and Relevance:** SICLI is an equitable, high-performing mortality prediction model that builds on and outperforms the EOLCI. It was implemented and pilot-tested in a large health care system.

**Key points:** *Question:* Can we build on a commercially available mortality risk score to develop, implement, and pilot-test a high-performing prediction model to support palliative care decision-making in a large integrated health care system?

*Findings:* The Serious Illness Clinical Indicator (SICLI) augments and outperforms the Epic End-of-Life Care Score (EOLCI) and was implemented and pilot-tested using a responsible AI framework.

*Meaning:* Predictive models can be valuable tools to support palliative care referrals.

## Introduction

Patients with serious illnesses are often referred late or not at all to palliative care,^1–3^ resulting in sub-optimal management of symptoms, pain and emotional distress as well as preventable emergency department visits, increased hospitalizations and a shorter time spent at home at the end of life.^4–9^ Predictive models have been proposed to support identification and timelier outreach to patients at the end of life.^10–14^ Numerous models have been developed, few have been implemented, and even fewer provide transparency across all stages of the model life cycle.^15,16^ Further, none of these models have been developed and implemented within a responsible artificial intelligence (AI) framework to assure fairness, transparency, accountability and trustworthiness of the prediction model.^17^ We follow TRIPOD-AI guidelines for transparent reporting and the Responsible AI Framework by the Organization of Economic Cooperation and Development (OECD) to inform the planning, development, implementation, validation and governance of a prediction tool to support decision making around palliative care.^18,19^

Prior models supporting palliative care clinical decision making have often been disease specific,^20–23^ or make use of very large datasets and sophisticated methods that render their implementation prohibitively complex for most health care systems.^24–27^ Epic, one of the largest electronic health record (EHR) providers in the United States,^28^ provides the Epic End-of-Life Care Index (EOLCI), a built-in regression based prediction model to assess 12-month mortality risk.^29^ The EOLCI was trained with data from three large, non-disclosed sites and relies on predictors that are available across most health care systems. Thus, it is highly portable but cannot take advantage of locally available data. Further, mortality models have been shown to generalize less well than expected to local health care systems.^30^

To build a more powerful prediction tool, we used an augment and tailor approach. This approach builds on, and adapts, the EOLCI to the specific needs and data availability of a large integrated health care system, Kaiser Permanente Southern California (KPSC), that serves a diverse population of 4.9 million members across 5 Southern California counties. Our approach augments the EOLCI with locally available data and optimizes model fit by training and validating the new model in the local population.^31,32^ This approach allows us to benchmark the performance of the new model against the EOLCI to decide if implementation of the new tool is warranted.

The KPSC Serious Illness Clinical Indicator (SICLI) predicts the risk of dying within 12 months among hospitalized patients at the 36^th^ hour of hospitalization. Its purpose was to support clinical decision making around the provision of primary and specialty palliative care as part of a large pragmatic trial.^33^ Physicians, informaticians and researchers collaborated closely in the planning, development, validation, implementation and testing of the model. Here we describe each stage of the model life cycle and the responsible AI principles that framed the development and implementation of SICLI in a large health care system.

## Methods

### Planning & Design

Our work follows the OECD framework for responsible AI which provides a comprehensive set of norms for each stage of the model life cycle and has been ratified by all OECD members (Figure 1).^18^ Responsible AI principles include 1) assurance of benefit to people of the AI model 2) human-centeredness of the tool to effectively assist decision making and assurance of statistical fairness of the model 3) transparency of model development, testing and implementation procedures 4) assurance of model robustness & data safety and 5) accountability for the implementation, monitoring and health care impact of the AI tool. We will describe below how we addressed each of these principles.

**Figure 1.**
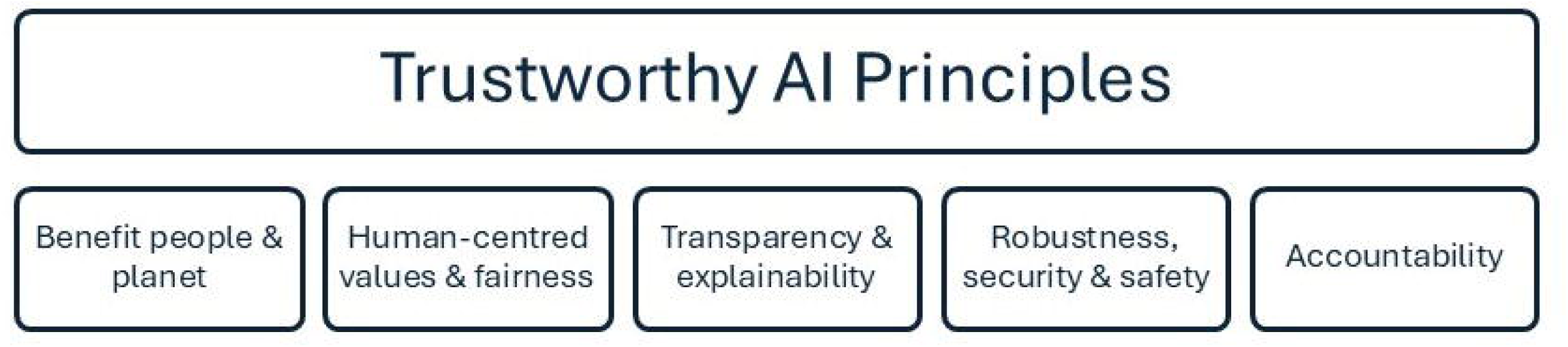
OECD responsible AI framework. This figure illustrates the five principles of the Organization for Economic Co-operation and Development (OECD) framework for responsible artificial intelligence, which guided the development and implementation of the SICLI model.

The purpose of SICLI was to support clinical decision making for hospitalized patients who may benefit from primary and/or specialty palliative care. Primary palliative care includes basic management of physical and emotional symptoms, advanced care planning and discussions of prognosis and goals of treatment by hospitalists and other subspecialists.^34^ Specialty palliative care is provided by board certified specialists and addresses complex symptom burden, care choices and family conflict that are more likely to occur in the later stages of serious illness.^34^ The benefit of identifying patients for primary and specialty care are multifold. Primary palliative care increases the likelihood of early introduction and continuity of palliative care services throughout the course of a patient’s serious illness and assures that the time of board-certified specialist palliative care clinicians can be focused on patients with the most complex needs.^34^

SICLI was designed to predict 12-month mortality to identify patients in need of palliative care and was trained to run at the 36^th^ hour of a patient’s hospitalization using data available at the time of the model run. The 36^th^ hour after admission was chosen based on stakeholder input as it allows sufficient time for a clinician to become familiar with a patient’s health status while also promoting earlier palliative care, consistent with observed evidence of its benefit.^35,36^ SICLI triggers an Epic “Our Practice Advisory” (OPAs) when a physician opens the patient chart for patients at high-risk (>=60 <95%) and very high-risk (>=95%) of mortality. The probability thresholds for high-risk (≥60- <95%) and very high-risk (≥95%) were selected by the research team in consultation with stakeholders to identify patients most likely to benefit from inpatient primary or specialty palliative care. Thresholds may be subject to change as the trial progresses.

This study was approved by the KPSC (13886) and University of Pennsylvania (855378) Institutional Review Boards.

### Training and validation cohort

SICLI was trained and validated on inpatient encounters from 15 KPSC hospitals from January 1^st^, 2022, to December 31^st^, 2022. Encounters of patients under the age of 18, those hospitalized in psychiatry and maternity departments or who died or were discharged before the 36^th^ hour were excluded. Data were split into a training dataset including the hospitalizations of 60% of patients (n of patients = 56,693, n of hospitalizations = 79,722). The encounters of the remaining 40% of patients (n of patients = 37,796, n of hospitalizations = 53,321) were used as a hold-out sample to validate the final model.

### Outcome

The outcome of interest was the predicted risk of dying within 12 months, a proxy for whether the patient was likely to need primary or specialty palliative care. Twelve-month prognosis has been commonly used as a proxy for the need for palliative care when using predictive models.^32,37^ Mortality data came from the EHR, membership data and the national death index.

### Candidate Predictor Set

The SICLI built on the EOLCI. The initial predictor set included 42 predictors documented in the EOLCI technical materials, such as 26 clinical classification system (CCS) diagnoses categories defined by the health care cost and utilization project, 15 medication categories, red cell distribution width, albumin, age, sex, and Medicaid status.^29^

To Augment the EOLCI, we expanded the predictor set to include the remaining 253 CCS disease categories and all active medication orders defined via therapeutic categories. Additionally, we included risk scores measuring overall health status and deterioration available in KPSC’s Epic instance. These included the Epic deterioration index, Laboratory and Acute Physiology Score, Version 2 (LAPS2), Comorbidity Point Score, Version 2 (COPS2),^38^ and the Sequential Organ Failure Assessment (SOFA) score.^39^ Based on clinical stakeholder input, we also included the glomerular filtration rate (GFR), information from nursing flow sheets, including the prior level of functioning at the time of admission (PLOF), coded as patient is able to walk >50 feet, walks < 50 feet, can stand, can sit, bed bound.

Health care utilization metrics were also added, including the number of emergency department visits and inpatient encounters in the prior six months and the source of admission (home vs nursing home, or another facility). The predictor set further included a serious illness indicator variable that was defined via an adapted list of diagnosis codes from the Center to Advance Palliative Care, as well as race/ethnicity categories (African American/Black, Asian, Hispanic/Latino, White and Other/unknown), to account for disparities in health outcomes and health care use.

We reviewed the predictor set with the clinical informatics team to exclude predictors that were unavailable or soon-to-be retired in the Epic environment. The final predictor set included 384 variables. To allow for easy implementation, missing values of non-categorical variables were median imputed. When a diagnosis was missing it was considered absent and set to 0. Variables with greater than 30% missingness were deleted.

### Model

KPSC’s Epic instance currently supports only regression-based models for real-time risk prediction. To meet this requirement, we used least absolute shrinkage and selection operator (LASSO) using logistic regression and five-fold cross-validation to predict 12-month mortality. The clinical informatics team confirmed that a model of up to 40 predictors was practical for implementation. We first identified the highest performing model (regardless of the number of predictors) based on the smallest cross-validated misclassification error. We then identified and compared model performance (AUC and calibration plots) of a model with the smallest classification error and up to 40 predictors. The final SICLI model was selected in collaboration with physicians and clinical informaticists who weighed performance and implementation burden.

### Algorithmic fairness assessment

Data availability and missingness were reviewed by population subgroups defined as African American, Asian, Hispanic, or of other/unknown race and ethnicity (eTable 1 in the Supplement). We compared model discrimination and calibration across patient groups. To ensure that each hospitalized patient who was dying had an equal chance of being identified by our model we further compared population group specific sensitivity.^40^ Where needed, sensitivity was improved by lowering group-specific probability thresholds.^41^ The resulting increase in referral volume and operational burden was assessed. Algorithmic equity was assessed for the high- and very-high risk group combined, as the sample size in the very-high risk group was small, and the goal was to ensure fair access to palliative care.

### Model documentation, model socialization and physician training

Model documentation for end-users supports model transparency and empowers clinicians to use the model effectively and safely for decision support.^42^ The research team and the chief of palliative care developed a document for clinicians that summarized the model’s purpose, performance and limitations (eFigure 1). SICLI was further socialized via departmental meetings with prospective physician end-users from hospital medicine and intensive care and other palliative care leaders.

### Post implementation evaluation

SICLI implementation was piloted at 3 KPSC hospitals from November 9 to December 20, 2024. We solicited feedback from hospitalists, intensivists, and palliativists within the first week, mid-point, and end of the 6-week pilot. To validate clinical performance of the model in real time (before the 12-month follow up period), a palliative care physician (S.E.W) reviewed 25 randomly selected charts (10 charts each in the low and high-risk groups and 5 charts in the very high-risk group) and assessed whether the patient was likely to die in the next 12 months.

## Results

Twelve-month mortality among 133,043 hospitalizations of at least 36 hours was 23% (Table 1). Patients who died within 12 months were older (76±13 vs. 64±17 years), had higher co-morbidity burden (Charlson, 4.49±3.14 vs. 2.06±2.44), were more likely to have a do-not-resuscitate/do not intubate order (33% vs. 9%), be bed-bound (18% vs. 5%) at admission, and have worse severity of hospitalization (Epic deterioration index: 39±15 vs. 29±11) compared to patients who did not die within 12 months. The training and validation samples were comparable (eTable 1).

**Table 1.** Descriptives of inpatient encounters.

| Characteristic | Full Data |  |  |
| --- | --- | --- | --- |
|  | Did not die<br>N = 102,602<sup>a</sup> | Died<br>N = 30,441<sup>a</sup> | Overall<br>N = 133,043<sup>a</sup> |
|  | 77,617 | 17,763 | 94,489 |
| **Age** | 64 (17) | 76 (13) | 67 (17) |
| <65 | 45,197 (44%) | 5,614 (18%) | 50,811 (38%) |
| >= 65 | 57,405 (56%) | 24,827 (82%) | 82,232 (62%) |
| **Gender** |  |  |  |
| Female | 52,884 (52%) | 14,508 (48%) | 67,392 (51%) |
| Male | 49,717 (48%) | 15,932 (52%) | 65,649 (49%) |
| Other | 1 (<0.1%) | 1 (<0.1%) | 2 (<0.1%) |
| **Race/Ethnicity** |  |  |  |
| Asian | 9,499 (9.3%) | 2,677 (8.8%) | 12,176 (9.2%) |
| Black | 13,232 (13%) | 4,528 (15%) | 17,760 (13%) |
| Hispanic | 38,847 (38%) | 9,506 (31%) | 48,353 (36%) |
| Other | 1,836 (1.8%) | 465 (1.5%) | 2,301 (1.7%) |
| White | 39,188 (38%) | 13,265 (44%) | 52,453 (39%) |
| **Insurance Status** |  |  |  |
| Medicaid/Dual-eligible | 14,202 (14%) | 3,990 (13%) | 18,192 (14%) |
| Non Medicaid | 88,400 (86%) | 26,451 (87%) | 114,851 (86%) |
| **Elixhauser Cancer** |  |  |  |
| Cancer | 22,144 (22%) | 13,240 (43%) | 35,384 (27%) |
| No Cancer | 80,458 (78%) | 17,201 (57%) | 97,659 (73%) |
| **Charlson Dementia** |  |  |  |
| Dementia | 7,893 (7.7%) | 6,578 (22%) | 14,471 (11%) |
| No Dementia | 94,709 (92%) | 23,863 (78%) | 118,572 (89%) |
| **Weighted Charlson Comorbidity Index** | 2.06 (2.44) | 4.49 (3.14) | 2.62 (2.81) |
| **Serious illness flag** | 56,532 (55%) | 28,469 (94%) | 85,001 (64%) |
| **CCS 42: Secondary Malignancies** | 3,840 (3.7%) | 5,428 (18%) | 9,268 (7.0%) |
| **CCS 252: Malaise and Fatigue** | 19,947 (19%) | 12,122 (40%) | 32,069 (24%) |
| **Last COPS 2.5 Score prior to 36<sup>th</sup> hour** | 46 (39) | 84 (50) | 55 (45) |
| **Last Epic-DI Score prior to 36<sup>th</sup> hour** | 29 (11) | 39 (15) | 31 (13) |
| **Last LAPS2 Score prior to 36<sup>th</sup> hour** | 56 (29) | 82 (38) | 62 (33) |
| **DNR code status at admission** | 9,043 (8.8%) | 9,922 (33%) | 18,965 (14%) |
| **RDW (most recent)** | 14.64 (2.35) | 16.25 (2.89) | 15.01 (2.57) |
| **PLOF (at admission) Bed Bound** | 5,023 (4.9%) | 5,390 (18%) | 10,413 (7.8%) |

<sup>a</sup> Mean (SD); n (%)
Abbreviations: CCS = Clinical Classifications Software, COPS = Comorbidity Point Score, Epic-DI = Epic Deterioration Index, LAPS2 = Laboratory-based Acute Physiology Score, DNR = Do not resuscitate, RDW = Red Cell Distribution Width test, PLOF = Prior Level of Function

The LASSO model with the lowest misclassification error had 325 variables and an AUC of 0.87 (95% CI: 0.87, 0.88). The EOLCI has a predictor set of 43 variables. The KPSC clinical informaticians team deemed a set of up to 40 predictors feasible to implement. Clinicians, clinical informaticians and researchers chose a parsimonious model with 39 variables that provided a high AUC (0.87, 95% CI: 0.86, 0.87) and excellent calibration (Figure 2, Panel A and Panel B). The full model including beta-coefficients is listed in Supplemental eTable 2. SICLI outperformed the EOLCI which produced an AUC of 0.81 (95% CI: 0.80, 0.81, Figure 2, Panel A) and notably under-predicted the mortality risk of high-risk patients (Figure 2, Panel B).

**Figure 2.**
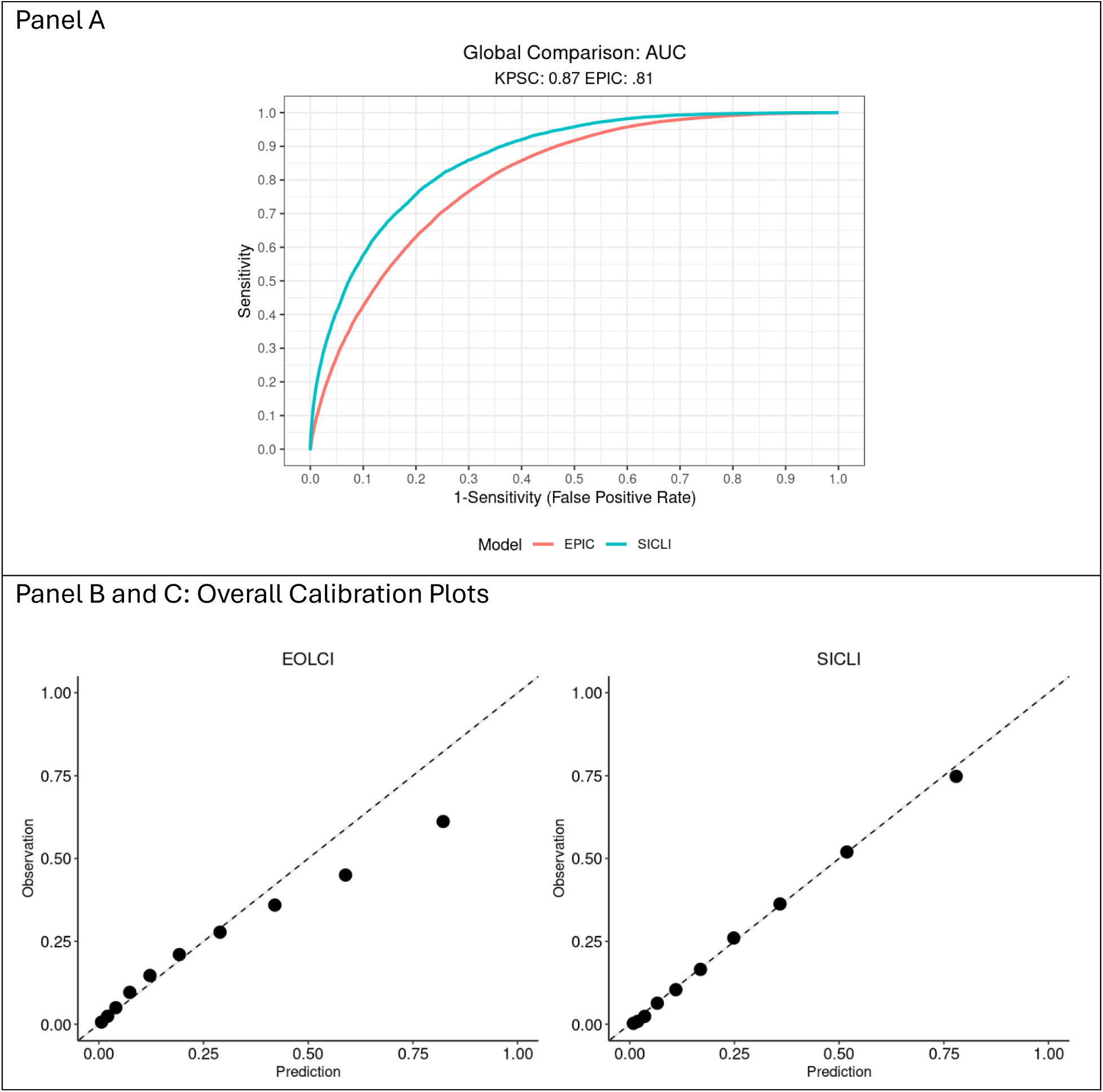
Discrimination and calibration of KPSC SICLI and Epic End-of-Life Care Index models. Panel A shows the area under the receiver operating characteristic curve (AUC) comparing the SICLI and Epic End-of-Life Care Index (EOLCI) models. Panel B displays calibration plots for both models, demonstrating the improved predictive accuracy and calibration of SICLI.

SICLI’s sensitivity at the 60% probability cut-off was 36% (combining high- and very high-risk groups). The positive predictive value (PPV) for the high- and very high-risk groups were 72.0% and 94.0%, respectively (Table 2). The low-risk group had a negative predictive value (NPV) of 83.68% and contained 88.88% of all hospitalizations (not shown). Algorithmic fairness assessment showed that the race/ethnicity specific AUCs were comparable, ranging from 0.85 for patients who were white to 0.88 for patients who were Asian, Hispanic and those included in the “other” category (eFigure 2). Population specific sensitivity was highest for patients who were Asian (40.6%) and lowest for patients in the other race/ethnicity category (28.8%). Lowering the probability cut-off for the other/race ethnicity category from 60% to 53.3% achieved a sensitivity that was comparable to the median sensitivity (34.7%). The adjustment of the probability cut-off did not affect the overall PPVs for the high and very-high risk groups.

**Table 2.** Group specific performance of SICLI.

|  | **High-risk category**<br>**($\geq 0.6$ - $\leq 0.94$)** | **Very high-risk category**<br>**($\geq 0.95$)** |
| --- | --- | --- |
| **Group specific PPV** | 72.05% | 94.03% |
| **Group specific sensitivity** | 33.21% | 2.48% |
| **Group Size**<br>**(annual number, percent of total)** | 5,552 (10.41%) | 299 (0.60%) |
| **False positives in risk group** | 1,552 | 19 |

**Table 3.** Equity assessment at a probability cut-off of 0.6 (before adjustment)

| **Race/Ethnicity** | **AUC (95% CI)** | **Sensitivity** |
| --- | --- | --- |
| **Asian** | 0.88 (0.87, 0.89) | 40.6% |
| **Black** | 0.86 (0.85, 0.87) | 38.9% |
| **Hispanic** | 0.88 (0.88, 0.89) | 34.4% |
| **Other** | 0.88 (0.85, 0.90) | 28.8% |
| **White** | 0.85 (0.84, 0.85) | 34.7% |
| **Median** | - | 34.7% |

Based on the 2022 cohort of hospitalized patients, SICLI would have triggered 14,794 alerts (11% of patient encounters) for patients with a 60% or greater risk of dying within 12 months. This would translate to an average of 2.7 alerts per day/hospital (2.56 and 0.14 for the high-risk and very high-risk groups, respectively). In comparison, the EOLCI would have triggered a total of 19,529 alerts (15% of patient encounters), an average of 3.57 alerts per day/hospital, 3.35 and 0.22 for the high and very-high risk group, respectively). Implementing the EOLCI instead of the SICLI would have created 4,735 additional alerts, of which 4,424 would have been false positives.

### Pilot performance

During the 6-week pilot, SICLI was computed for 3,848 eligible hospitalizations and identified 517 patients with a predicted risk of >=60%. Chart reviews within 2-5 months of the hospitalization confirmed that all 5 patients in the very high-risk group were likely to die within the next 12 months, yielding a chart review-based PPV of 100% compared to a PPV of 94% when calculated on the SICLI risk-prediction for the 2022 cohort. Three of the five patients in the very high-risk group had died at the time of chart review. Among the 10 patients identified as high risk, the palliativist considered six to be likely to die within 12 months. The chart review-based PPV for the high-risk group was 80.0% compared to the 72.0% PPV calculated based on the model predictions.

### Responsible AI principles

Our approach followed the OECD framework for responsible AI, which emphasizes benefit to people, human-centeredness, transparency, robustness, and accountability (Figure 1). ^18^ Pilot performance was promising and we continue to monitor SICLI’s performance and health care impact to assure its benefit to patients (first principle). SICLI was designed to be a human-centered AI tool (second principle) that provides decision support and care recommendations to the physician who remains the key decision maker (human in the loop).^18^ Algorithmic fairness has been assessed (second principle) and remediated to assure equal benefit for all patients. To assure transparency (third principle), we developed context-appropriate model documentation for physicians and the detailed description of all stages of the model life cycle provided herein. Model robustness and safety (fourth principle) was assured by developing a parsimonious model following the TRIPOD AI guidelines. Data safety is addressed by storing individual risk scores in a separate EHR data table, with access limited to IRB approved staff tasked with monitoring the performance of SICLI. To avoid the tool being used for other purposes or by users not trained in its use, the patient’s mortality risk category is not recorded in the patient chart. Finally, accountability for the risk score has been assumed by the research team, the chief of palliative care, the medical informatics team, and KPSC’s Systems Solutions and Deployment team who assumed oversight of the deployment and continued monitoring of SICLI’s model performance and impact on care.

## Discussion

SICLI is an equitable, high-performing, and well-calibrated mortality prediction model designed to support clinical decision making around primary and specialty palliative care for hospitalized patients. We used an augment and tailor approach to build on and improve the performance of a widely used Epic prediction tool (EOLCI). We described how SICLI was planned, developed, validated, implemented and piloted, following both transparent reporting standards (TRIPOD AI), and responsible AI principles.^18,19^

Most prior work on clinical decision support for palliative care has focused on model development.^10,21,43,44^ However, models implemented in the inpatient setting show promise; a randomized controlled trial demonstrated an increase in palliative care referrals and reduction in 60- and 90-day re-admission rates.^17^ Another pilot study showed increases in palliative care consultations, advanced care planning and reduction in the length of stay.^11^ A rule-based algorithm to identify patients for referrals to palliative care improved palliative consultation rates and increased the rates of discharge to hospice and do not resuscitate orders.^45^

SICLI built on these promising results and showed further promise during pilot testing. Continuous monitoring of SICLI’s performance ensures its ongoing effectiveness. While algorithmic equity was assessed and improved, equitable use of a prediction model further requires culturally appropriate communication about palliative care services. KPSC’s chief of palliative care has implemented extensive serious illness communication training and cultural humility training for physicians that supports the use of SICLI.

While the development of SICLI followed best practice guidelines, our approach has important limitations. Firstly, we did not assess if model performance could be enhanced with other machine learning models such as deep learning or extreme gradient boosting because the KPSC Epic instance currently does not support the implementation of such models. We tailored the Epic EOLCI to the KPSC context to improve its performance. This results in our tool being less generalizable to other sites than the EOLCI. Our approach to augmenting and tailoring the EOLCI can be adopted by other health care systems and is likely to improve model performance through in-sample training and increasing the predictor set.

SICLI attempted to balance parsimony, practicability and responsiveness to local context and was successfully implemented and piloted. We offer a road map to a pragmatic approach for improving existing risk scores and responsibly implementing the resulting decision support tool.

## Supporting information

SICLI Supplement

## Author Contributions

**Conceptualization:** CN, SEW, HQN

**Methodology:** CN, SEW, MH, HZ, BH, HQN

**Formal analysis:** CN, BH, HZ, MH

**Investigation:** CN, SEW, MH, HZ, LV, SMS, KM, KD, BH, BC, SH, KC, JSL, HQN

**Data curation:** CN, HZ, MH, JSL

**Writing – original draft:** CN

**Writing – review & editing:** All authors

**Supervision:** HQN

**Project administration:** BC

CN had full access to all the data in the study and takes responsibility for the integrity of the data and the accuracy of the data analysis.

## Data Sharing Statement

The data are available for use through collaboration with the KPSC study investigators under the conditions of sufficient funding by the requestor and data use agreements between all institutions that govern data access, storage, and short- and long-term use. Qualified researchers trained in human subject confidentiality protocols interested in collaborating with the KPSC study team can contact Dr. Claudia Nau.

## References

1. Iyer AS, Sullivan DR, Lindell KO, Reinke LF. The Role of Palliative Care in COPD. Chest. 2022;161(5):1250–1262. doi:10.1016/j.chest.2021.10.032

2. Kayastha N, LeBlanc TW. When to Integrate Palliative Care in the Trajectory of Cancer Care. Curr Treat Options Oncol. 2020;21(5):41. doi:10.1007/s11864-020-00743-x

3. Chang YK, Kaplan H, Geng Y, et al. Referral Criteria to Palliative Care for Patients With Heart Failure. Circ Heart Fail. 2020;13(9):e006881. doi:10.1161/CIRCHEARTFAILURE.120.006881

4. Bekelman DB, Feser W, Morgan B, et al. Nurse and Social Worker Palliative Telecare Team and Quality of Life in Patients With COPD, Heart Failure, or Interstitial Lung Disease: The ADAPT Randomized Clinical Trial. JAMA. 2024;331(3):212–223. doi:10.1001/jama.2023.24035

5. Quinn KL, Shurrab M, Gitau K, et al. Association of Receipt of Palliative Care Interventions With Health Care Use, Quality of Life, and Symptom Burden Among Adults With Chronic Noncancer Illness: A Systematic Review and Meta-analysis. JAMA. 2020;324(14):1439–1450. doi:10.1001/jama.2020.14205

6. Kavalieratos D, Corbelli J, Zhang D, et al. Association Between Palliative Care and Patient and Caregiver Outcomes: A Systematic Review and Meta-analysis. JAMA. 2016;316(20):2104–2114. doi:10.1001/jama.2016.16840

7. Gonzalez-Jaramillo V, Fuhrer V, Gonzalez-Jaramillo N, Kopp-Heim D, Eychmüller S, Maessen M. Impact of home-based palliative care on health care costs and hospital use: A systematic review. Palliat Support Care. 2021;19(4):474–487. doi:10.1017/S1478951520001315

8. Yadav S, Heller IW, Schaefer N, et al. The health care cost of palliative care for cancer patients: a systematic review. Support Care Cancer. 2020;28(10):4561–4573. doi:10.1007/s00520-020-05512-y

9. Wang SE, Gozansky WS, Steiner C, et al. Association Between Intensity and Timing of Specialty Palliative Care and Hospice Exposure With Quality of End-of-Life Care. J Palliat Med. 2024;27(5):602–613. doi:10.1089/jpm.2023.0407

10. Wang L, Sha L, Lakin JR, et al. Development and Validation of a Deep Learning Algorithm for Mortality Prediction in Selecting Patients With Dementia for Earlier Palliative Care Interventions. JAMA Netw Open. 2019;2(7):e196972. doi:10.1001/jamanetworkopen.2019.6972

11. Courtright KR, Chivers C, Becker M, et al. Electronic Health Record Mortality Prediction Model for Targeted Palliative Care Among Hospitalized Medical Patients: a Pilot Quasi-experimental Study. J Gen Intern Med. 2019;34(9):1841–1847. doi:10.1007/s11606-019-05169-2

12. Murphree DH, Wilson PM, Asai SW, et al. Improving the delivery of palliative care through predictive modeling and healthcare informatics. J Am Med Inform Assoc. 2021;28(6):1065–1073. doi:10.1093/jamia/ocaa211

13. Reddy V, Nafees A, Raman S. Recent advances in artificial intelligence applications for supportive and palliative care in cancer patients. Curr Opin Support Palliat Care. 2023;17(2):125. doi:10.1097/SPC.0000000000000645

14. Wilson PM, Ramar P, Philpot LM, et al. Effect of an Artificial Intelligence Decision Support Tool on Palliative Care Referral in Hospitalized Patients: A Randomized Clinical Trial. J Pain Symptom Manage. 2023;66(1):24–32. doi:10.1016/j.jpainsymman.2023.02.317

15. Bressler T, Song J, Kamalumpundi V, Chae S, Song H, Tark A. Leveraging Artificial Intelligence/Machine Learning Models to Identify Potential Palliative Care Beneficiaries: A Systematic Review. J Gerontol Nurs. 2025;51(1):7–14. doi:10.3928/00989134-20241210-01

16. Rakers MM, van Buchem MM, Kucenko S, et al. Availability of Evidence for Predictive Machine Learning Algorithms in Primary Care: A Systematic Review. JAMA Netw Open. 2024;7(9):e2432990. doi:10.1001/jamanetworkopen.2024.32990

17. Kuziemsky CE, Chrimes D, Minshall S, Mannerow M, Lau F. AI Quality Standards in Health Care: Rapid Umbrella Review. J Med Internet Res. 2024;26:e54705. doi:10.2196/54705

18. Organization for Economic Cooperation and Development. Advancing Accountability in AI: Governing and Managing Risks throughout the Lifecycle for Trustworthy AI.; 2023. Accessed March 21, 2025. 10.1787/2448f04b-en

19. Collins GS, Moons KGM, Dhiman P, et al. TRIPOD+AI statement: updated guidance for reporting clinical prediction models that use regression or machine learning methods. BMJ. Published online April 16, 2024:e078378. doi:10.1136/bmj-2023-078378

20. Gensheimer MF, Teuteberg W, Patel MI, et al. Automated patient selection and care coaches to increase advance care planning for patients with cancer. JNCI J Natl Cancer Inst. 2025;117(2):296–302. doi:10.1093/jnci/djae243

21. Cary MP, Zhuang F, Draelos RL, et al. Machine Learning Algorithms to Predict Mortality and Allocate Palliative Care for Older Patients with Hip Fracture. J Am Med Dir Assoc. 2021;22(2):291–296. doi:10.1016/j.jamda.2020.09.025

22. Khera R, Haimovich J, Hurley NC, et al. Use of Machine Learning Models to Predict Death After Acute Myocardial Infarction. JAMA Cardiol. 2021;6(6):633–641. doi:10.1001/jamacardio.2021.0122

23. Norgeot B, Glicksberg BS, Trupin L, et al. Assessment of a Deep Learning Model Based on Electronic Health Record Data to Forecast Clinical Outcomes in Patients With Rheumatoid Arthritis. JAMA Netw Open. 2019;2(3):e190606. doi:10.1001/jamanetworkopen.2019.0606

24. Chi S, Guo A, Heard K, et al. Development and Structure of an Accurate Machine Learning Algorithm to Predict Inpatient Mortality and Hospice Outcomes in the Coronavirus Disease 2019 Era. Med Care. 2022;60(5):381. doi:10.1097/MLR.0000000000001699

25. Avati A, Jung K, Harman S, Downing L, Ng A, Shah NH. Improving palliative care with deep learning. BMC Med Inform Decis Mak. 2018;18(4):122. doi:10.1186/s12911-018-0677-8

26. Wang L, Sha L, Lakin JR, et al. Development and Validation of a Deep Learning Algorithm for Mortality Prediction in Selecting Patients With Dementia for Earlier Palliative Care Interventions. JAMA Netw Open. 2019;2(7):e196972. doi:10.1001/jamanetworkopen.2019.6972

27. Vu E, Steinmann N, Schröder C, et al. Applications of Machine Learning in Palliative Care: A Systematic Review. Cancers. 2023;15(5):1596. doi:10.3390/cancers15051596

28. Bruce G. Epic’s dominance in 12 numbers. Becker’s Hospital Review | Healthcare News & Analysis. July 18, 2024. Accessed March 21, 2025. https://www.beckershospitalreview.com/ehrs/epics-dominance-in-12-numbers/

29. Epic. Cognitive Computing Model Brief: End of Life Care Index. Epic; 2023.

30. Singh H, Mhasawade V, Chunara R. Generalizability challenges of mortality risk prediction models: A retrospective analysis on a multi-center database. PLOS Digit Health. 2022;1(4):e0000023. doi:10.1371/journal.pdig.0000023

31. Nau C, Butler RK, Huang CW, et al. Development and Validation of the COVID-19 Hospitalized Patient Deterioration Index. Am J Manag Care. 2023;29(12):e365–e371. doi:10.37765/ajmc.2023.89470

32. Gensheimer MF, Lu J, Ramchandran K. Comparison of 1-year mortality predictions from vendor-supplied versus academic model for cancer patients. PeerJ. 2025;13:e18958. doi:10.7717/peerj.18958

33. Halpern SD, Courtright K. Comparative Effectiveness of Generalist versus Specialist Palliative Care for Inpatients | PCORI. Patient-Centered Outcomes Research Institute. July 18, 2023. Accessed May 9, 2025. https://www.pcori.org/research-results/2023/comparative-effectiveness-generalist-versus-specialist-palliative-care-inpatients

34. Ernecoff NC, Check D, Bannon M, et al. Comparing Specialty and Primary Palliative Care Interventions: Analysis of a Systematic Review. J Palliat Med. 2020;23(3):389–396. doi:10.1089/jpm.2019.0349

35. May P, Normand C, Cassel JB, et al. Economics of Palliative Care for Hospitalized Adults With Serious Illness: A Meta-analysis. JAMA Intern Med. 2018;178(6):820–829. doi:10.1001/jamainternmed.2018.0750

36. May P, Garrido MM, Cassel JB, et al. Prospective Cohort Study of Hospital Palliative Care Teams for Inpatients With Advanced Cancer: Earlier Consultation Is Associated With Larger Cost-Saving Effect. J Clin Oncol. 2015;33(25):2745–2752. doi:10.1200/JCO.2014.60.2334

37. Gupta A, Burgess R, Drozd M, Gierula J, Witte K, Straw S. The Surprise Question and clinician-predicted prognosis: systematic review and meta-analysis. BMJ Support Palliat Care. 2025;15(1):12–35. doi:10.1136/spcare-2024-004879

38. Escobar GJ, Gardner MN, Greene JD, Draper D, Kipnis P. Risk-adjusting hospital mortality using a comprehensive electronic record in an integrated health care delivery system. Med Care. 2013;51(5):446–453. doi:10.1097/MLR.0b013e3182881c8e

39. Vincent JL, Moreno R, Takala J, et al. The SOFA (Sepsis-related Organ Failure Assessment) score to describe organ dysfunction/failure. Intensive Care Med. 1996;22(7):707–710. doi:10.1007/BF01709751

40. Rajkomar A, Hardt M, Howell MD, Corrado G, Chin MH. Ensuring Fairness in Machine Learning to Advance Health Equity. Ann Intern Med. 2018;169(12):866–872. doi:10.7326/M18-1990

41. Paulus JK, Kent DM. Predictably unequal: understanding and addressing concerns that algorithmic clinical prediction may increase health disparities. Npj Digit Med. 2020;3(1):99. doi:10.1038/s41746-020-0304-9

42. Sendak MP, Gao M, Brajer N, Balu S. Presenting machine learning model information to clinical end users with model facts labels. Npj Digit Med. 2020;3(1):41. doi:10.1038/s41746-020-0253-3

43. Aixia Guo, Foraker R, White P, Chivers C, Courtright K, Moore N. Using Electronic Health Records and Claims Data to Identify High-risk Patients Likely to Benefit From Palliative Care. Am J Manag Care. 2021;27(1):e7–e15. doi:10.37765/ajmc.2021.88578

44. Owusuaa C, van der Padt-Pruijsten A, Drooger JC, et al. Development of a Clinical Prediction Model for 1-Year Mortality in Patients With Advanced Cancer. JAMA Netw Open. 2022;5(11):e2244350. doi:10.1001/jamanetworkopen.2022.44350

45. Courtright KR, Madden V, Bayes B, et al. Default Palliative Care Consultation for Seriously Ill Hospitalized Patients: A Pragmatic Cluster Randomized Trial. JAMA. 2024;331(3):224–232. doi:10.1001/jama.2023.25092

