## Supplementary material for "Responsible AI in Action: Planning through Implementation of a Mortality Model for Palliative Care": SICLI Supplement

**Online Supplement**

### eTable 1. Proportion of missing median-imputed variables, by race/ethnicity

| Characteristic | Asian | Black | Hispanic | Multiple | Native Am/ Alaskan | Other | Pacific Islander | Unknown | White | Overall |
| --- | --- | --- | --- | --- | --- | --- | --- | --- | --- | --- |
|  | N = 12,176*^a^* | N = 17,760*^a^* | N = 48,353*^a^* | N = 340*^a^* | N = 263*^a^* | N = 619*^a^* | N = 892*^a^* | N = 187*^a^* | N = 52,453*^a^* | N = 133,043*^a^* |
| GFR | 14 (0.1%) | 24 (0.1%) | 76 (0.2%) | 1 (0.3%) | 1 (0.4%) | 1 (0.2%) | 1 (0.1%) | 1 (0.5%) | 60 (0.1%) | 179 (0.1%) |
| RDW | 2 (<0.1%) | 9 (<0.1%) | 46 (<0.1%) | 1 (0.3%) | 1 (0.4%) | 0 (0%) | 1 (0.1%) | 0 (0%) | 56 (0.1%) | 116 (<0.1%) |
| Last LAPS2 Score  prior to 36th hour | 0 (0%) | 0 (0%) | 0 (0%) | 0 (0%) | 0 (0%) | 0 (0%) | 0 (0%) | 0 (0%) | 0 (0%) | 0 (0%) |
| Last COPS2.5 Score prior to 36th hour | 602  (4.9%) | 1,701  (9.6%) | 4,309  (8.9%) | 17  (5.0%) | 24  (9.1%) | 48  (7.8%) | 69  (7.7%) | 20  (11%) | 4,761  (9.1%) | 11,551  (8.7%) |
| Last IP Deterioration  Index Prior to 36th hour | 0 (0%) | 4 (<0.1%) | 3 (<0.1%) | 0 (0%) | 0 (0%) | 0 (0%) | 0 (0%) | 0 (0%) | 4 (<0.1%) | 11 (<0.1%) |
| New Score prior to 36th hour | 0 (0%) | 0 (0%) | 0 (0%) | 0 (0%) | 0 (0%) | 0 (0%) | 0 (0%) | 0 (0%) | 0 (0%) | 0 (0%) |

*^a^*n (%)

### eTable 2. Implemented model variables and coefficients (not standardized)

| Variable | Estimate |
| --- | --- |
| (Intercept) | -7.928231919 |
| Modified Charlson Index | 0.023292811 |
| Epic Deterioration Index | 0.021682848 |
| Cops2 | 0.000815709 |
| Serious Illness Indicator | 1.175738435 |
| Laps2 | 0.008516136 |
| Full Code/Any Other Code Status | 0.612906293 |
| RDW | 0.103997523 |
| Age | 0.026674413 |
| CCS_42, Malignant Neoplasm Wo Specification Of Site | 0.817009037 |
| Ccs_252, Malaise And Fatigue | 0.067744043 |
| Bed Bound Indicator | 0.701202143 |
| CCS_52, Nutritional Deficiencies | 0.090469788 |
| CCS_45, Maintenance Chemotherapy/Radiotherapy | 0.460432564 |
| Ccs_L1_2 Neoplasms | -0.006480316 |
| Dementia Indicator | 0.358754112 |
| CCS_155, Other Gastrointestinal Disorders | 0.104485805 |
| CCS_17, Cancer Of Pancreas | 1.168121904 |
| CCS_130 Pleurisy; Pneumothorax; Pulmonary Collapse | 0.186378272 |
| CCS_258 Other Screening For Suspected Conditions (Not Mental Disorders Or Infectious Disease) | 0.205701245 |
| Cancer Indicator | 0.460203545 |
| CCS_16, Cancer Of Liver And Intrahepatic Bile Duct | 0.855632588 |
| SOFA Score | 0.041577861 |
| Can Walk < 50FT Indicator | 0.370941334 |
| CCS_55 Fluid And Electrolyte Disorders | 0.087244013 |
| Count Of IP Stays In Last 6 Months | 0.039661632 |
| CCS_10 Immunizations And Screening For Infectious Disease | 0.044093810 |
| CCS_255 Administrative/Social Admission | 0.073007151 |
| Mild So To Severe Liver Disease Indicator | 0.417127981 |
| CCS_43 Malignant Neoplasm Without Specification Of Site | 0.625920584 |
| CCS_35 Cancer Of Brain And Nervous System | 1.237300603 |
| CCS_19 Cancer Of Bronchus; Lung | 0.365149900 |
| CCS_151 Other Liver Diseases | 0.130831304 |
| Therapeutic Class, Colony Stimulating Factors | 0.139723675 |
| Therapeutic Class, CNS | -0.240704124 |
| Able To Sit Indicator | 0.507802654 |
| Admission Source Group Home Or Clinic | -0.181940869 |
| CCS_62 Coagulation And Hemorrhagic Disorders | 0.030590030 |
| Ccs_247 Lymphadenitis | 0.331174104 |
| Can Stand Indicator | 0.532306375 |

### eFigure 1. Brief model documentation for end-users

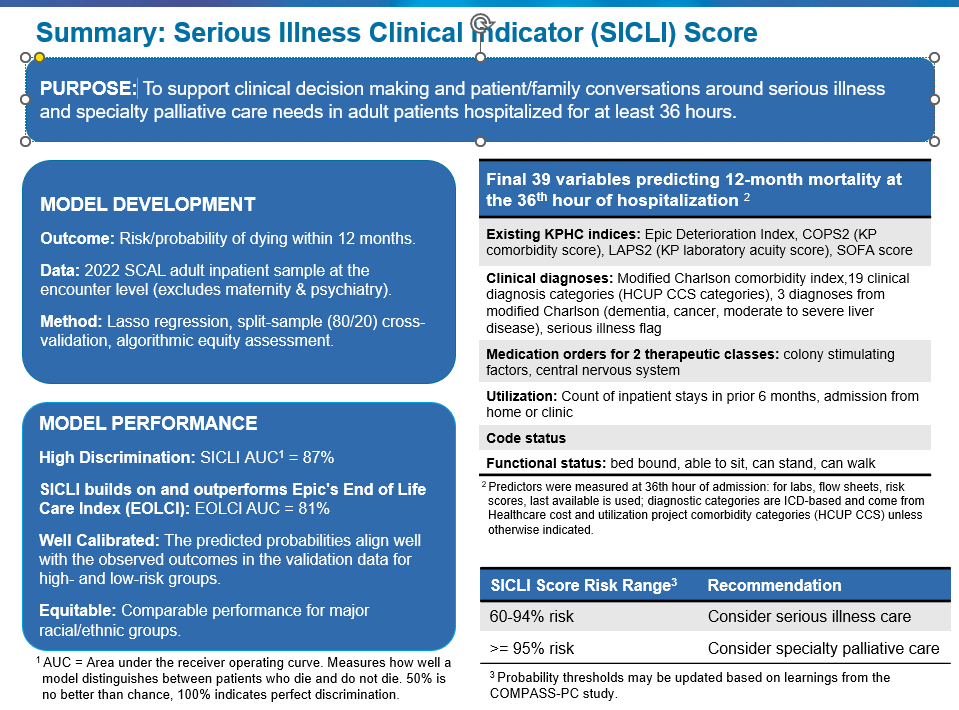

eFigure 2. Calibration plots by Race/Ethnicity

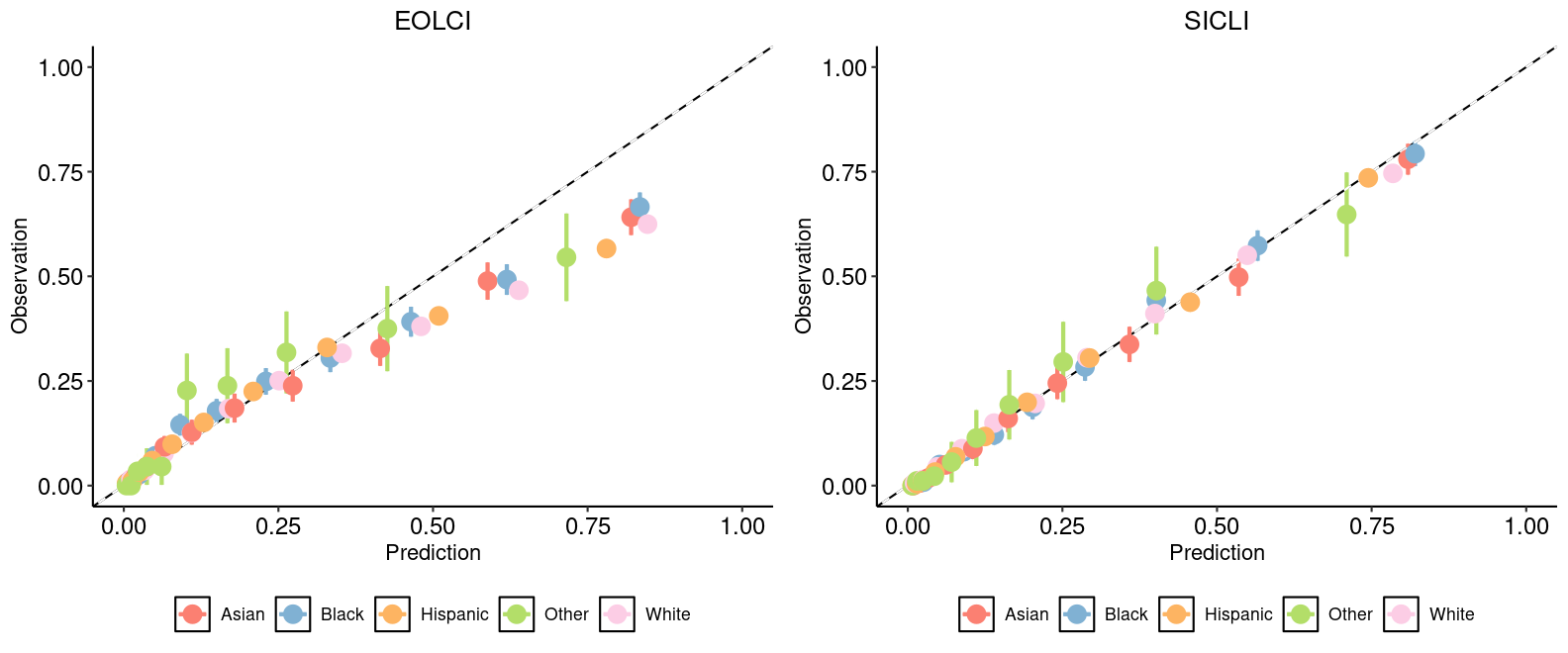
